# Racial/Ethnic Disparities in Youth Mental Health Traits and Diagnoses within a Community-based Sample

**DOI:** 10.1101/2023.02.13.23285862

**Authors:** Andrew Dissanayake, Annie Dupuis, Christie Burton, Noam Soreni, Paul Peters, Amy Gajaria, Paul D. Arnold, Jennifer Crosbie, Russell Schachar

## Abstract

**Background:** Racial/ethnic disparities in the prevalence of mental health diagnoses have been reported but have not accounted for the prevalence of the traits that underlies these disorders. Examining rates of diagnoses in relation to traits may yield a clearer understanding of how racial/ethnic youth differ in their access to assessment and care. We sought to examine differences in self/parent-reported rates of diagnoses for obsessive-compulsive disorder (OCD), attention-deficit/ hyperactivity disorder (ADHD), and anxiety disorders after adjusting for differences in trait levels between youth from three racial/ethnic groups: White, South Asian, and East Asian.

**Methods:** We collected parent or self-reported ratings of OCD, ADHD and anxiety traits and diagnoses for youth (6-17 years) from a general population sample (Spit for Science). We examined racial/ethnic differences in trait levels and the odds of reporting a diagnosis using mixed-effects linear models and logistic regression models.

**Results:** East Asian (N = 1301) and South Asian (N = 730) youth reported significantly higher levels of OCD and anxiety traits than White youth (N = 6896). Given the same trait level, East Asian and South Asian youth had significantly lower odds of reporting a diagnosis for OCD (Odds Ratio (OR) _East Asian_ = 0.08 [0.02, 0.41] ; OR _South Asian_ = 0.05 [0.00, 0.81]), ADHD (OR _East Asian_ = 0.27 [0.16, 0.45]; OR _South Asian_ = 0.09 [0.03, 0.30]), and Anxiety (OR _East Asian_ = 0.21 [0.11, 0.39]; OR _South Asian_ = 0.12 [0.05, 0.32]) than White youth.

**Conclusions:** These results suggest a discrepancy between traits-levels of OCD and anxiety and rates of diagnoses for East Asian and South Asian youth. This discrepancy may be due to increased barriers for ethnically diverse youth to access mental health care. Efforts to understand racial/ethnic barriers to care are needed.

**Key Points:**

- Despite having lower prevalence of diagnoses, East and South Asian youth reported significantly higher anxiety and OCD trait levels than White youth
- Given the same trait level, East Asian youth were at 92% lower odds of having received an OCD diagnosis, 73% lower odds of having received an ADHD diagnosis, and 79% lower odds of having received an Anxiety diagnosis than White youth
- Given the same trait level, South Asian youth were at 95% lower odds of having received an OCD diagnosis, 91% lower odds of having received an ADHD diagnosis, and 88% lower odds of having received an anxiety diagnosis
- Future research is needed to understand barriers to mental health care and assessment that may underly the discrepancy between mental health traits and diagnoses for ethnic/racially diverse youth.

## INTRODUCTION

Ethnic and racial disparities in health status and access to care are present in many domains of health care (Mahajan et al., 2021). These disparities are especially salient in the field of mental health (Alegria, Vallas, & Pumariega, 2010; Chow, Jaffee, & Snowden, 2003; Coker et al., 2016; Cook, Nhi-Ha, Zhihui, Sherry Shu-Yeu, & Ana, 2017) warranting calls for increased scientific attention (Haeny, Holmes, & Williams, 2021). Some studies suggest that ethnic and racial minorities have a lower prevalence of several psychiatric disorders (Breslau, Kendler, Su, Gaxiola-Aguilar, & Kessler, 2005; Chiu, Amartey, Wang, & Kurdyak, 2018; Chung et al., 2019; Coker et al., 2016). However, there is also evidence that suggests ethnic and racial minorities have worse mental health (Chiu et al., 2018; Werner, 2020) but are more likely to face significant barriers to mental health assessment and care resulting in unmet needs (Chiu et al., 2018; Tiwari & Wang, 2008). These mixed findings may arise because large population-based studies examine rates of physician-diagnosed mental health diagnoses (Asnaani, Richey, Dimaite, Hinton, & Hofmann, 2010) without examining differences in the underlying prevalence of symptoms or traits of a mental illness between different ethnic groups within the general population.

Race and ethnicity are social constructs (as opposed to biological constructs) that can act as significant socio-political lenses through which racism and inequality can be understood (Flanagin, Frey, Christiansen, & AMA Manual of Style Committee, 2021). Though distinct constructs, there is considerable overlap between the general use and understanding of the terms “ethnicity” and “race”. Ethnicity refers to a person’s cultural identity (eg, language, customs, religion) whereas race refers to broad categories of people that are divided arbitrarily based on ancestral origin and physical characteristics (Bhopal, 2004; Flanagin et al., 2021; Flores, 2020). There has been increasing concern about what has been deemed an arbitrary separation of ethnicity and race in equity-based research (Flanagin et al., 2021; Flores, 2020). Proposals have been put forth to treat race and ethnicity as non-mutually exclusive categories of a single concept. Henceforward, we refer to race/ethnicity as a single variable.

The mechanisms by which race/ethnicity affects mental health care remain largely unclear. Some studies suggest affect access due to reduced willingness to engage in mental health (Chiu et al., 2018; Tiwari & Wang, 2008) though the factors underlying this reduced willingness willingness to engage in mental health services must be barriers to accessing care such as a lack of culturally competent, options. Other explanations suggest that the quality of mental health services may differ by race /ethnicity (U.S. Department of Health and Human Services, 1999) and that can be affected by experiences of discrimination, exposure to discrimination and stigma (Bailey et al., 2017; Eylem et al., 2020; Pal, Sharan, & Chadda, 2017) within the community (Shim et al., 2012). Racial/ethnic identity has also been shown to have both positive and negative associations with the prevalence of mental health traits and symptoms depending on the disorders and different racial/ethnic groups examined (Ching & Williams, 2019; Sundquist, 1993; J. J. Wang, Lin, Best, Selles, & Stewart, 2020). It remains unclear whether differential rates of mental health diagnoses between ethnic groups reflect differences in the population prevalence of mental health symptoms.

Social, cultural, and economic risk factors may also contribute to youth’s mental health and youth access to mental health care by influencing the accessibility of services and quality of services available (Macintyre, Ferris, Gonçalves, & Quinn, 2018). In understanding access to treatment, prevalence rates, and outcomes, it is important to consider socioeconomic variables to ensure that indicators such as unemployment, poverty, and poor housing, which may be independently linked with poor mental health, are not the primary drivers for differences in rates (Silva, Loureiro, & Cardoso, 2016). This is particularly relevant as racially and ethnically diverse young people often have an increased prevalence of these social risk factors due to historical discrimination and racism. Without controlling for the contribution of economic adversity, adverse outcomes may be falsely attributed to race and ethnicity when in fact they are driven by structural factors. Controlling for economic adversity is important in the Canadian context as although health care is publicly funded, many mental health services are not.

Another limitation of the existing literature examining mental health disparities is that it is largely based on adult populations (Chiu et al., 2018; Chow et al., 2003; Chung et al., 2019; Walker, Cummings, Hockenberry, & Druss, 2015) and samples based in the United States (Coker et al., 2016; Cook et al., 2017; Mahajan et al., 2021; Morgan, Staff, Hillemeier, Farkas, & Maczuga, 2013; Shi et al., 2021) where healthcare is private. In Canada, the degree to which these disparities are present among Canadian youth within a largely publicly funded healthcare system is unclear.

In order to better appreciate the landscape of mental health difficulties for racially and ethnically diverse Canadian young people, we must better understand the prevalence of mental illnesses within the population. In particular, given the hypothesized differential access to care, it is important to estimate prevalence using non-clinical samples which include individuals who may not otherwise have access to clinical care. This is a complex topic, and estimations need to also take into account the multiple forces that affect mental health in racially and ethnically diverse youth (e.g., socioeconomic status). In order to answer this question, this study sought to examine racial/ethnic differences in levels of self- and parent-reported mental health traits and the prevalence of reported mental health diagnoses (while adjusting for differences in trait levels), within a large Canadian community-based sample of children and youth. We tested the following primary research questions: 1) Do youth from specific racial/ethnic groups have different levels of anxiety, obsessive-compulsive disorder (OCD) and attention-deficit/hyperactivity disorder (ADHD) traits; and 2) Given the same trait level, do youth from specific minority racial/ethnic groups have lower odds of reporting a mental health diagnosis and treatment for anxiety, OCD, and ADHD than white youth. As a secondary aim, we examined whether social and economic factors mediated reported differences in mental health trait levels and/or rates of diagnoses and treatment between racial/ethnic groups.

## METHODS

### Participants

Participants were part of the Spit for Science study and were recruited while visiting a science museum (Ontario Science Center) between 2008 and 2009. For the purposes of this study, we only included participants residing in Canada. A demographic breakdown of the entire sample (N = 16062) is available in Table S1. Parents provided ratings for younger children (n = 13162 Mean age = 10.2 ± 2.2). Youths ages 12 and older who were able to provide informed consent, reported on their own trait levels and diagnoses (n = 2900, Mean age = 15.3 ± 1.4) . Further details about the study and study participants are available in supplemental materials.

### Ethical Considerations

We obtained informed consent, and verbal assent where applicable, for all participants as approved by the Hospital for Sick Children Research Ethics Board.

### Measures

#### Race/Ethnicity

Race/Ethnicity was measured by self- or parent-reported race/ethnicity of the participant’s maternal and paternal grandparents. Race/ethnicity responses were coded in accordance with the Canadian Institute for Health’s Proposed Standards for Race-Based and Indigenous Identity Data Collection and Health Reporting in Canada (Canadian Institute for Health Information, 2020). We used a threshold of all four grandparents of a given race/ethnicity for a participant to be classified as that given race/ethnicity. For our analyses, we retained participants who were classified as White, East Asian and South Asian, as these were the only race/ethnic groups sufficiently large to be analyzed. Although we wished to include mixed participants in our analyses, participants with multiple different combinations of entries made it difficult to separate the participants into meaningful subgroups with sufficient sample sizes. However, with hopes of including more mixed participants in our analyses, we also conducted multiple sensitivity analyses where the race/ethnicity definition threshold was altered (ie. >2 grandparents of X race/ethnicity = X race/ethnicity) and observed no major differences in conclusions or effect size estimates.

#### Trait Measures

Mental health traits are characteristics that are continuously distributed within the general population. There has been increasing evidence suggesting that ADHD, OCD, and Anxiety disorders are extremes of a continuous trait (Burton et al., 2019; Park et al., 2016). The distribution of these quantitative traits in the general population is relevant to the study of racial/ethnic disparities in mental health because traits may act as a proxy for the underlying distribution of a given mental illness within a given population. ADHD trait levels were measured by the Strengths and Weaknesses of Attention-Deficit/Hyperactivity-symptoms and Normal-behaviors (SWAN) rating scale (Burton et al., 2019). OCD symptoms were measured by The Toronto Obsessive-Compulsive Scale (TOCS) (Park et al., 2016). Anxiety and trait levels were measured by the anxiety subscale within the child behaviour checklist (CBCL) (Knepley, Kendall, & Carper, 2019). Measurement invariance between each racial/ethnic group included in this study for each measure was established at the scalar level enabling mean comparisons between groups (Putnick & Bornstein, 2016). We calculated t-scores for each symptom standardized for the respondent, age, and gender (when significantly associated with the raw scores). To examine the discrepancy between individuals with high trait levels likely warranting a diagnosis and the rates of reported diagnoses/treatment, we classified individuals as being “high-trait” based on the estimated population prevalence of a given mental health disorder. High trait for OCD was defined as having OCD traits scores in the 99th percentile of the population. High trait for ADHD was defined as having greater than the 95th percentile for either inattentive, hyperactive, or overall trait score. Anxiety high trait was defined as having trait scores greater than the 97.5th percentile within the population.

#### Reported Diagnoses

Reported diagnoses for ADHD, OCD, and Anxiety were measured by participants’ response to the question: “Have you had a diagnosis of, or treatment for any of the items listed below” with a list of mental health disorders to select from.

#### Socioeconomic Status and Neighborhood Marginalization Indices

For a subgroup of participants residing in the Greater Toronto Area (the region around the Ontario Science Centre), herein labelled as the “Urban subgroup”, we estimated socioeconomic status based on the first three digits of participants’ Postal Code by calculating the quintile of neighbourhood income per person equivalent, adjusted for household size, which is released by Statistics Canada. We further obtained established neighbourhood-level indices of marginalization including material deprivation, residential instability, dependency, and ethnic concentration based on the participant’s Postal Code (Matheson, Dunn, Smith, Moineddin, & Glazier, 2012). Material deprivation is defined as “refers to the inability for individuals or households to afford those consumption goods and activities that are typical in a society at a given point in time, irrespective of people’s preferences with respect to these items” (Matheson et al., 2012). Residential instability refers to “when the frequency of residential mobility in a household or individual is high or occurs over short intervals (Leventhal & Newman, 2010; Matheson et al., 2012). Dependency refers to “area-level concentrations of people who don’t have income from employment” (Matheson et al., 2012). Ethnic concentration refers to “area-level concentrations of recent immigrants and people belonging to a visible minority group” (Matheson et al., 2012). We only estimated these indices within the subgroup of participants residing in the Greater Toronto Area to ensure data quality in terms of spatial variability and using 3-digit postal codes for residential proxy is primarily valid for those in urban areas.

#### Statistical Analyses

To examine differences in ADHD, OCD and Anxiety trait levels between ethnic groups, we estimated multiple mixed-effect linear models where standardized t-scores for each symptom were the dependent variables. Race/Ethnicity was a categorical predictor, with White youth treated as the reference group given that it was the largest category. Sibling-relatedness was accounted for as a random effect in these models (Eu-ahsunthornwattana et al., 2014). To examine the possibility that results differed by respondent, we re-ran the models with the raw score (unadjusted for respondent) stratified by respondent.

To examine ethnic differences in rates of mental health diagnosis, we described the proportion of individuals who reported a diagnosis and/or treatment for the given mental illnesses. To examine differences in the rates of mental health diagnosis after adjusting for participant symptom levels between ethnic and racial groups, we fit Firth binomial logistic regression models accounting for rare events(Firth, 1993; X. Wang, 2014) with ethnicity and race as a categorical predictor and reported diagnosis for a given mental illness as the dependent variable. We then use the model’s regression coefficients to calculate adjusted odds ratios for each ethnic and racial group in comparison to White youth. Sibling relatedness was accounted for in the logistic regression models by randomly retaining one participant per family. All models were covariate-adjusted for age and gender. We fit subsequent models adjusting for covariates such as socioeconomic status, primary language spoken at home, and additional indices of marginalization: residential instability, material deprivation, dependency, and ethnic concentration in the subgroup of participants residing in GTA. We also compared the prevalence of individuals with “high traits” for a given disorder with the prevalence of reported diagnoses for each ethnic and racial group.

## RESULTS

### Participant Characteristics

Of those with complete ethnicity and race, mental health symptoms, and diagnosis data, 7244 were classified as White, 1379 were classified as East Asian, and 790 were classified as South Asian. The South Asian (mean age =11.3) and East Asian mean age = 11.1) youth were older than the White youth (mean age=10.7) (Table 1). 54.6 % of the sample resided in the Greater Toronto Area. Consistent with the younger age among White participants, the proportion of parent-reported data as opposed to self-reported data was higher in the group who were classified as White (Table 1).

**Table 1:**
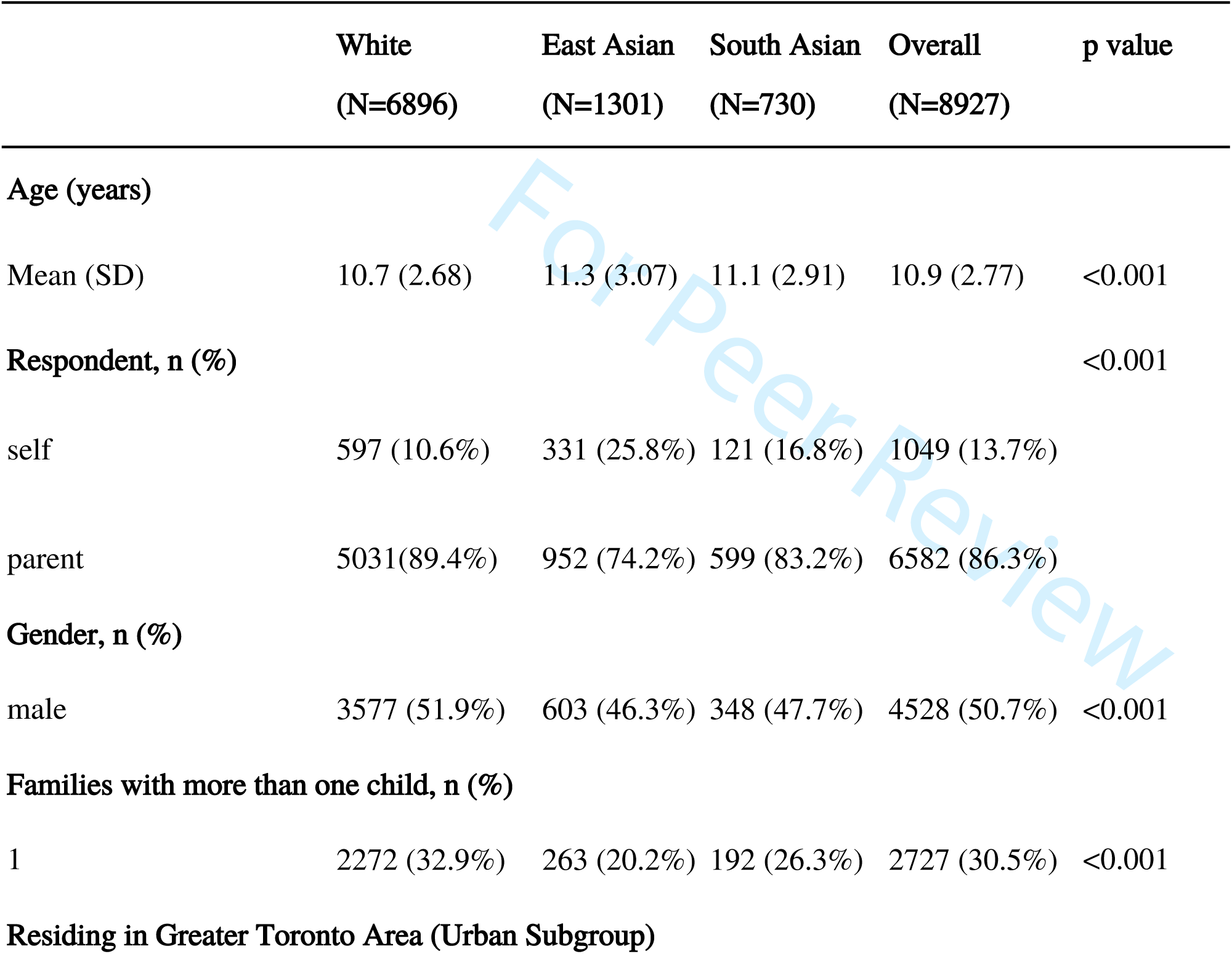
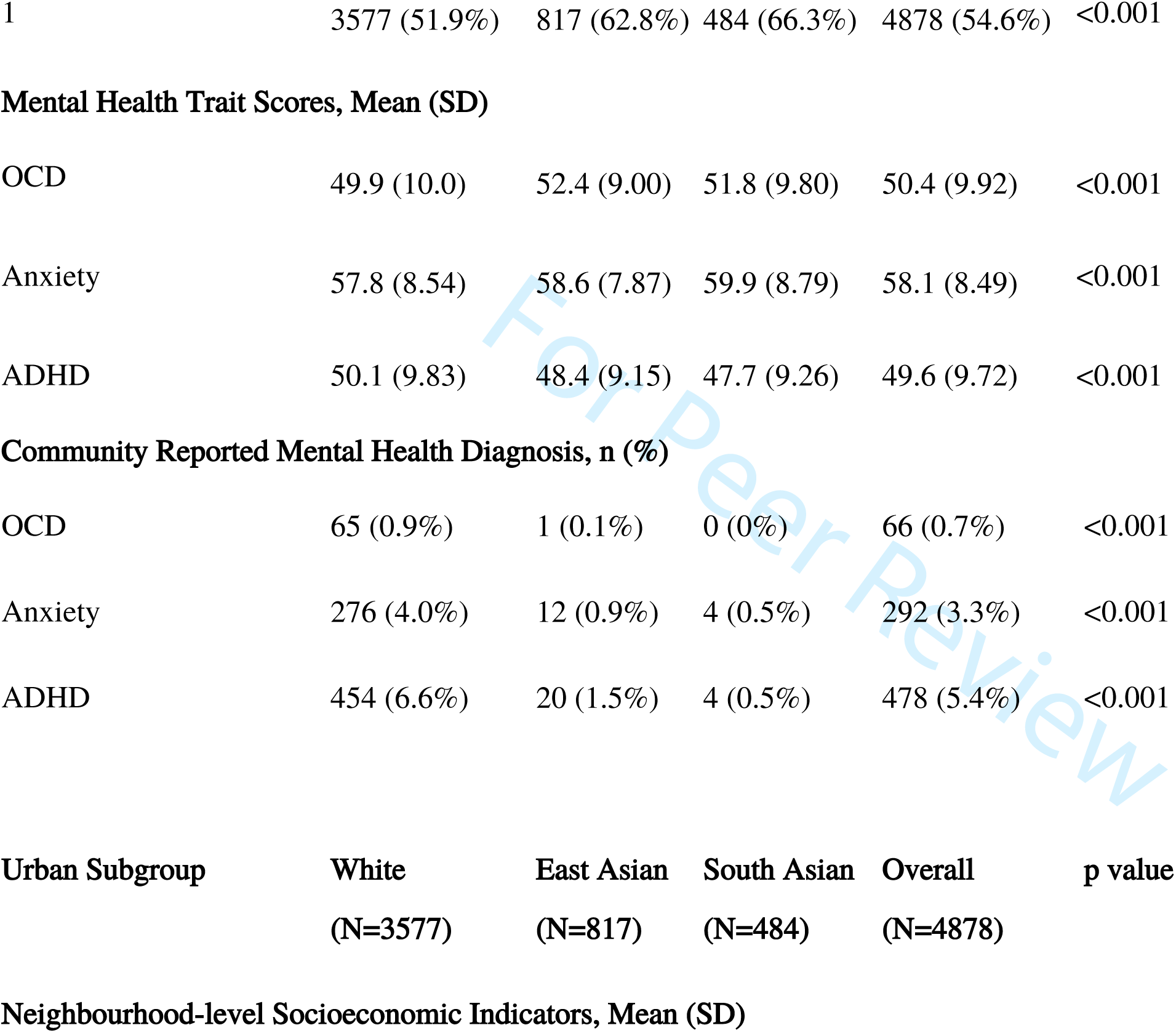
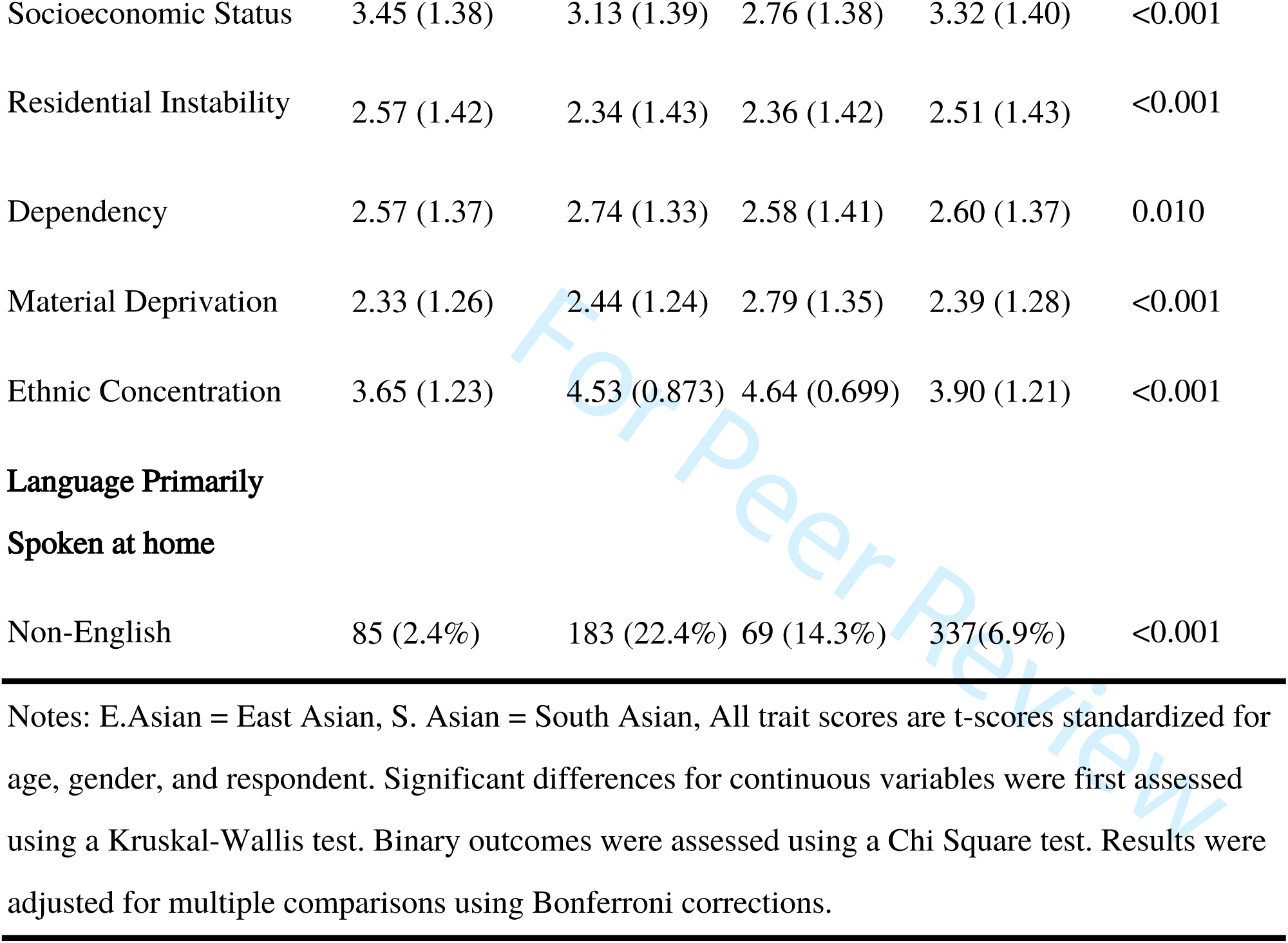
Sample Characteristics

### Ethnic Differences in Neighbourhood Marginalization Levels

Within the subsample of participants residing in the Greater Toronto Area, East Asian and South Asian participants were of significantly lower socioeconomic status than White participants (Table 1). South Asian and East Asian youth, on average, lived in neighbourhoods with significantly higher levels of ethnic concentration and material deprivation (Table 1). On average, East Asian youth lived in neighbourhoods with higher levels of dependency than White youth (Table 1). White youth tended to live in neighbourhoods with significantly higher levels of residential instability than East Asian and South Asian youth (Table 1).

### Racial/ethnic Differences in Mental Health Trait Levels

East Asian and South Asian youth had significantly higher levels of OCD traits and anxiety traits compared to White youth. (Table 2). However, East Asian and South Asian youth had significantly lower levels of ADHD traits. Results in the total sample were consistent in the Greater Toronto Area subsample even after adjustment for covariates. Stratification by the respondent did not Results in the sample of the Urban subgroup, were consistent for OCD and ADHD, even after adjusting for covariates including age, gender, material deprivation, ethnic concentration, dependency, residential instability, and socioeconomic status (Table 2). We also observed that socioeconomic status was inversely associated with ADHD trait levels and that material deprivation was positively associated with ADHD traits. Further, we found that ethnic concentration was associated with increased anxiety trait levels (Table 2).

**Table 2:**
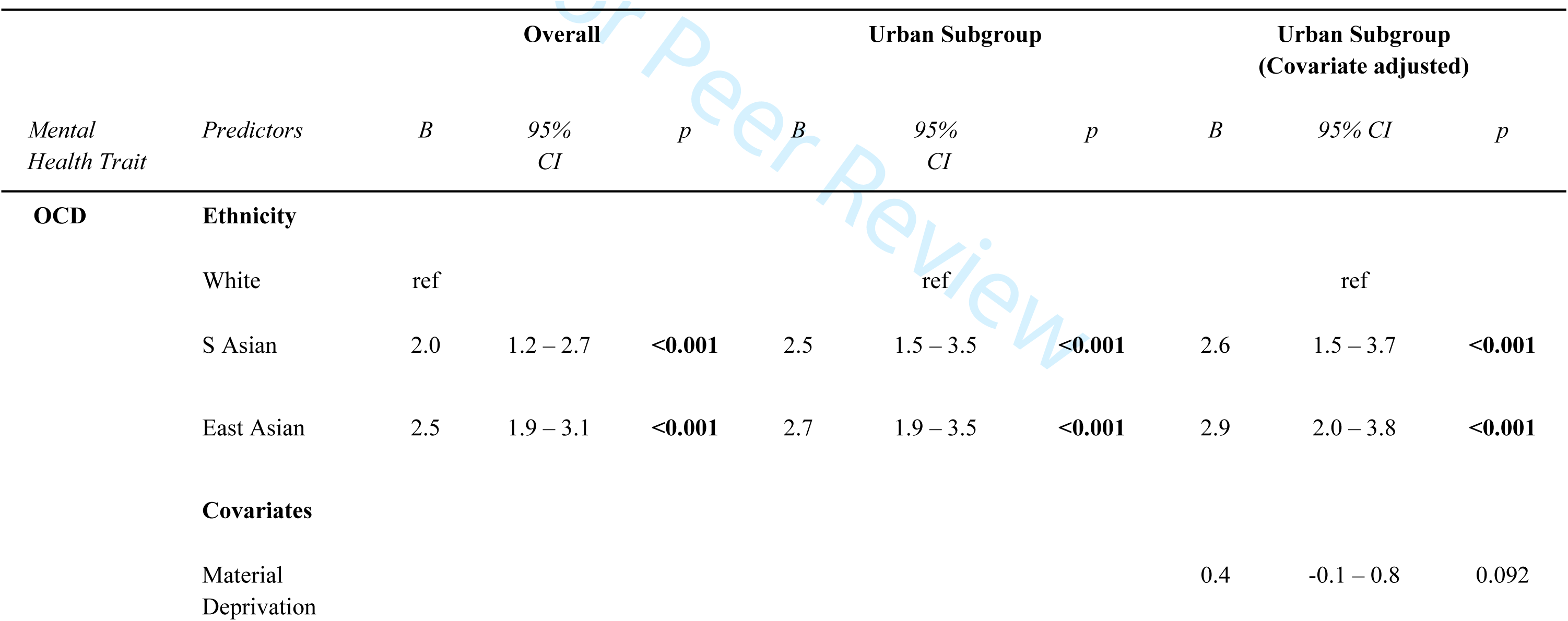
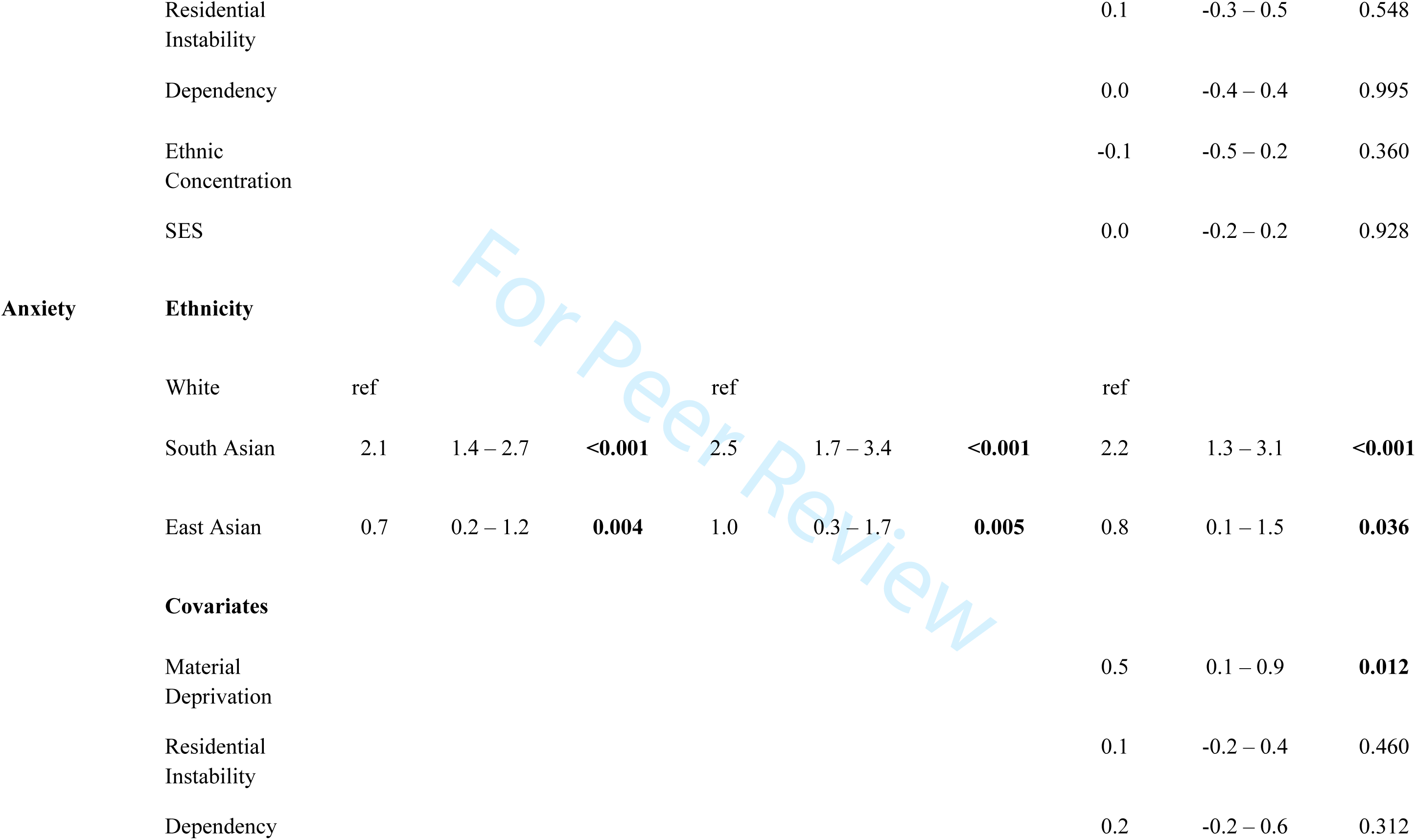
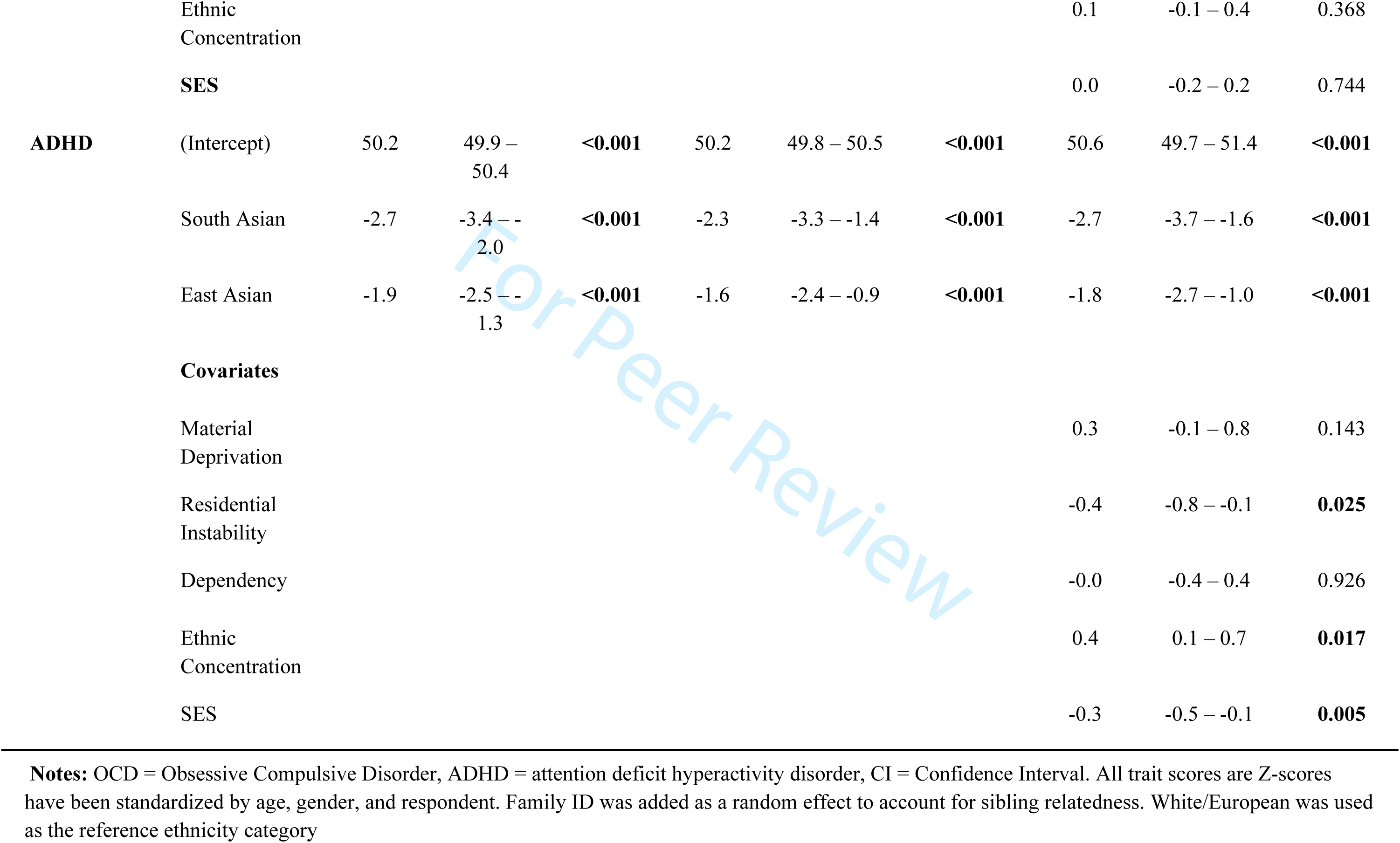
Racial/Ethnic Differences in Mental Health Trait Levels

### Ethnic Differences in Odds of Reporting Diagnosis

Given the same OCD trait level, South Asian youth had 95% decreased odds of reporting a diagnosis and/or treatment for OCD compared to White youth (Table 3). Similarly, East Asian youth had 77% lower odds of reporting a diagnosis and/or treatment for OCD (Table 3). South Asian and East Asian youth had 86% and 76% lower odds, respectively, of reporting a diagnosis and/or treatment for Anxiety compared to White youth (Table 3). For ADHD, we found that given the same trait level, South Asian and East Asian youth had 88% and 75% lower odds of reporting a diagnosis or treatment for ADHD compared to White youth (Table 3). All results showed no major differences in the [XXXXXXX] subsample, after adjusting for covariates with similar effect sizes and p-values. Increasing age was significantly associated with increased odds of reporting a diagnosis for each disorder examined. Females were significantly less likely to report a diagnosis/treatment for ADHD given the same trait level, but no gender differences were found for Anxiety and OCD. Having a non-english language primarily spoken at home was associated with lower odds of reporting a diagnosis and/or treatment for Anxiety (Table 3)

**Table 3:**
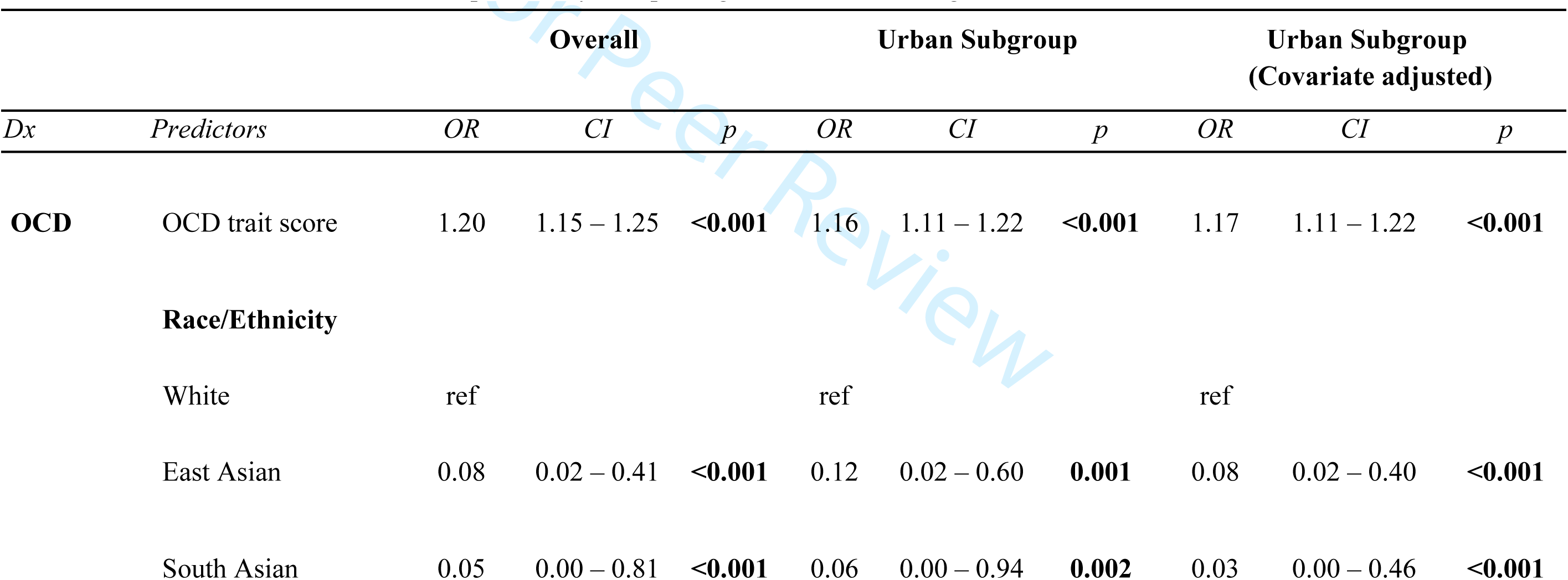
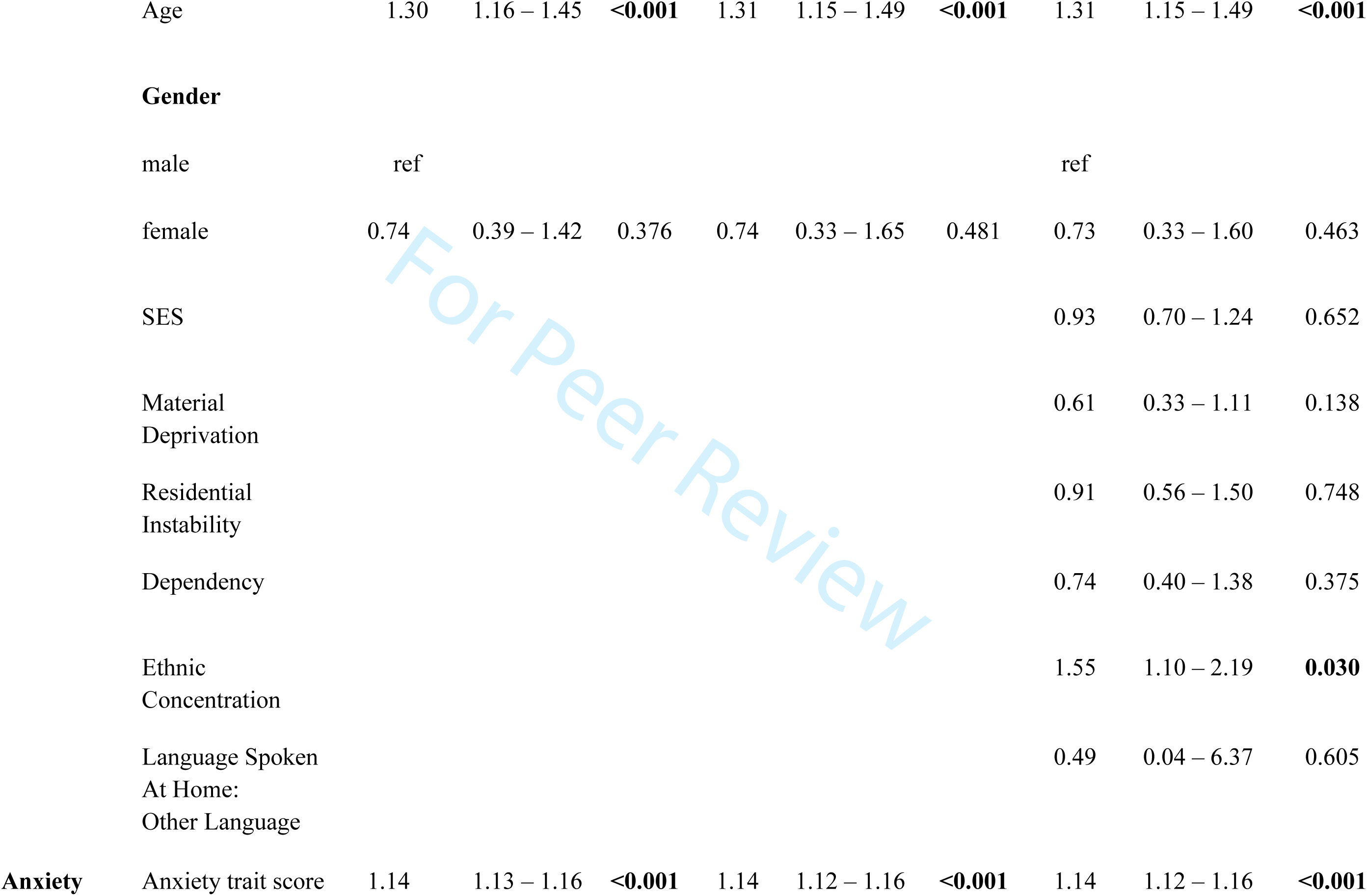
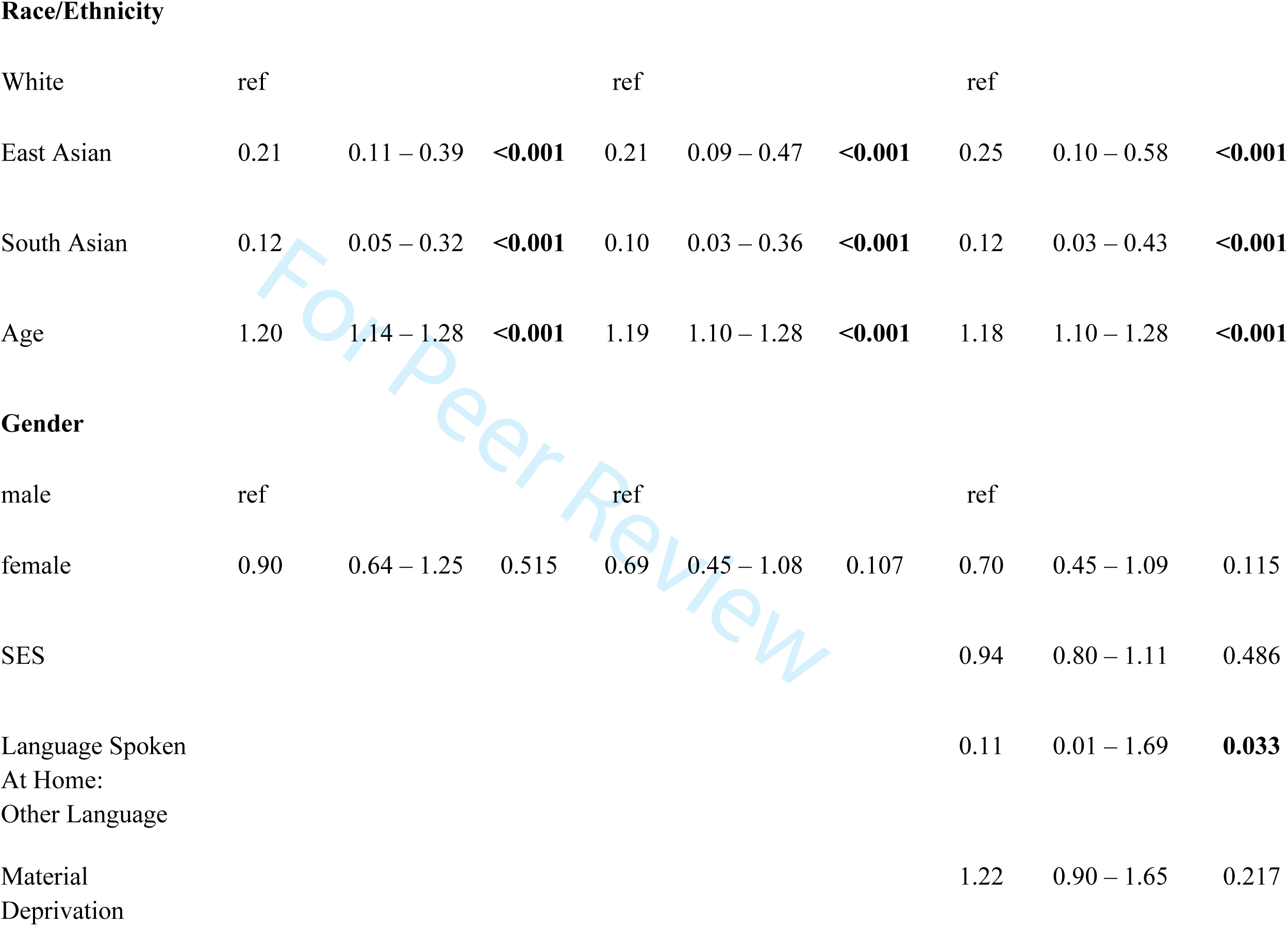
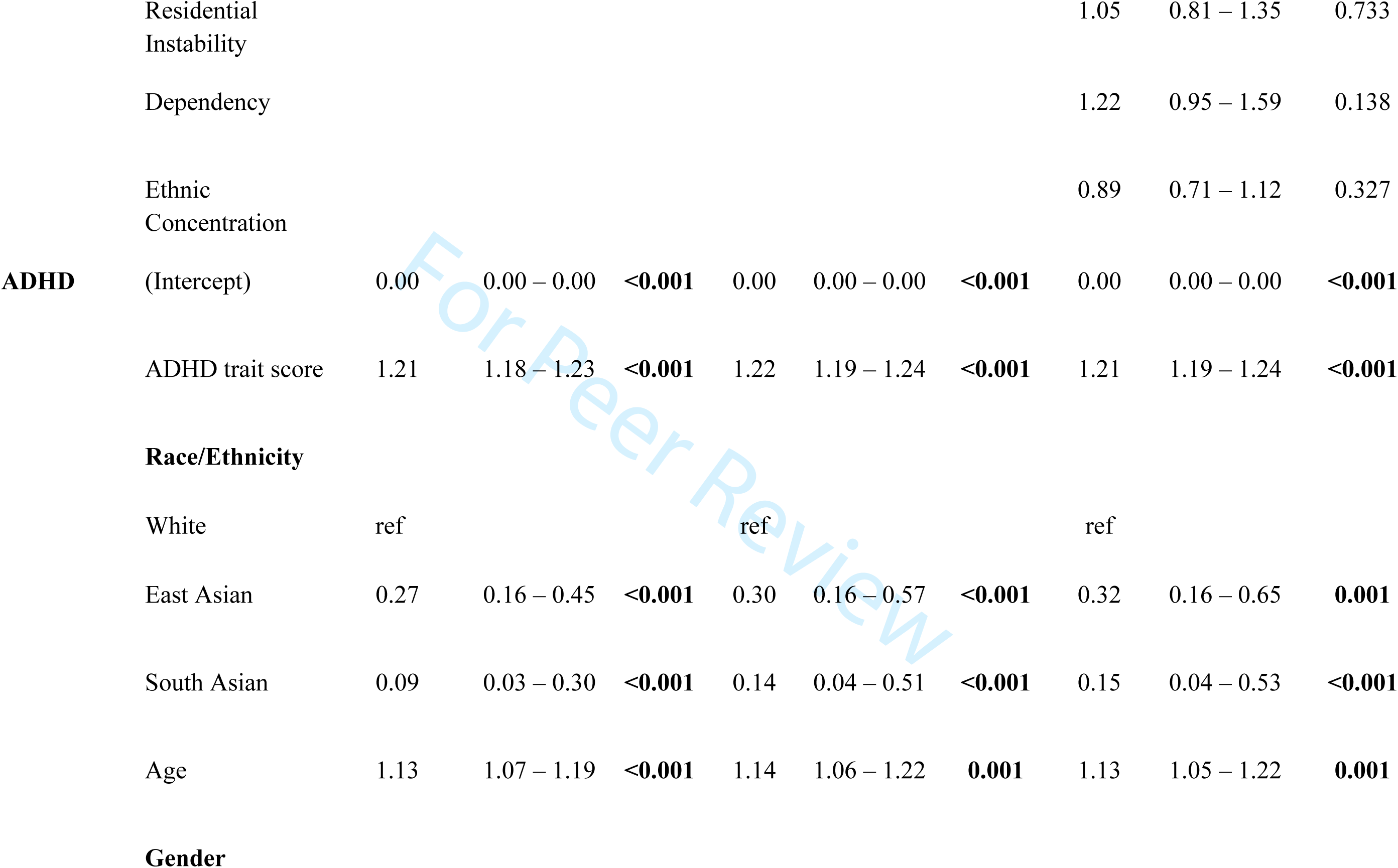
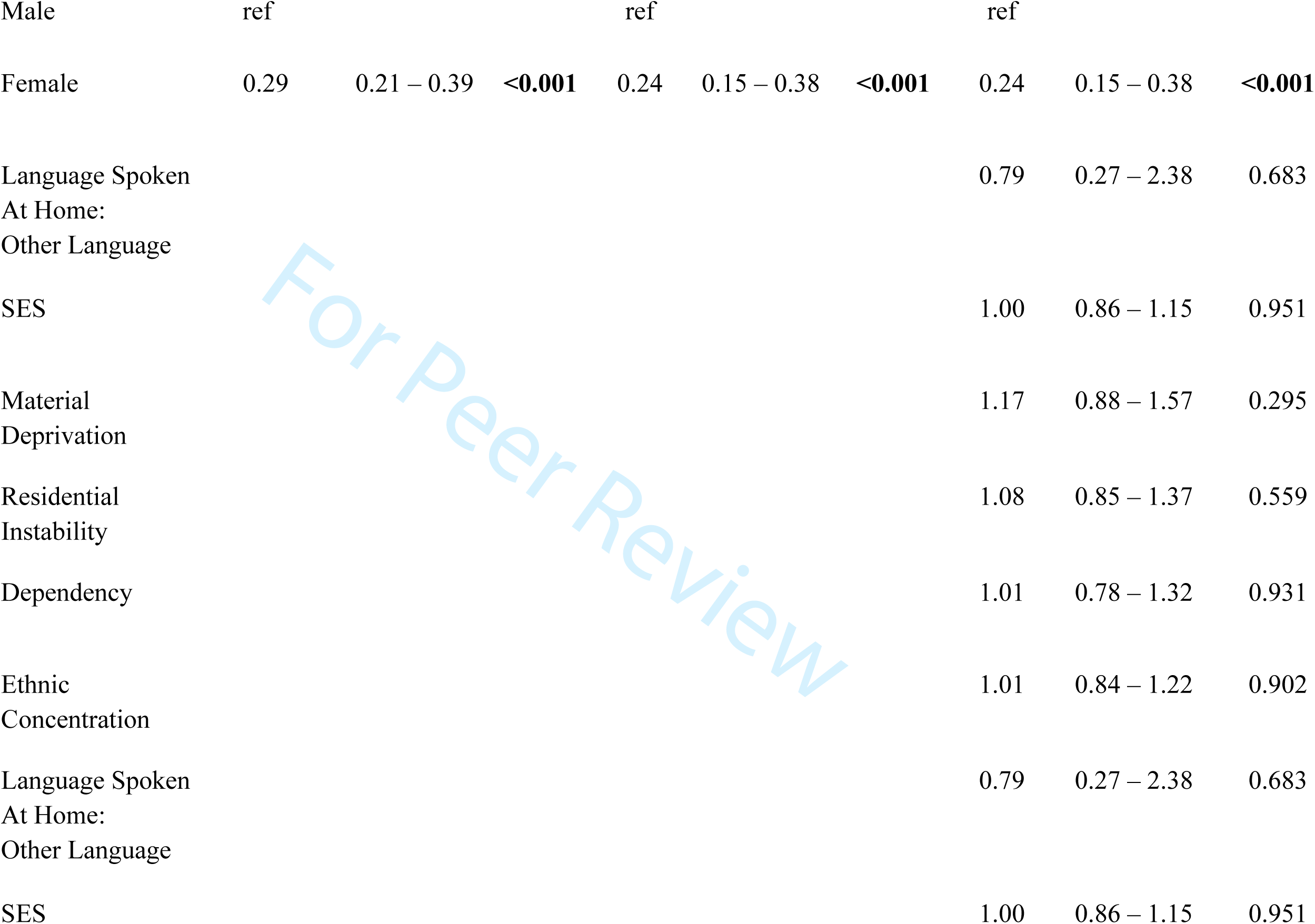
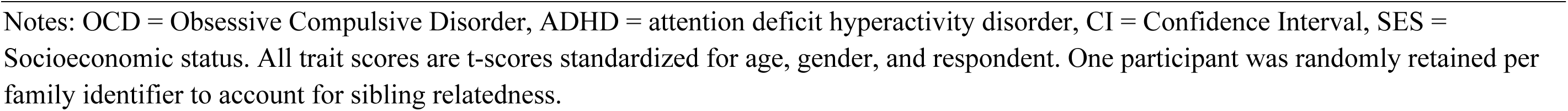
Racial/Ethnic Differences in the probability of reporting a mental health diagnosis

### Examining the Prevalence of High trait levels versus Community Diagnosis between Racial/ethnic Groups

We further compared the proportion of individuals with high trait levels of a given condition with the proportion who had reported a diagnosis (Figure 2). For OCD, Anxiety, and ADHD, there was a discrepancy between the prevalence of high trait levels versus the prevalence of those who received a diagnosis within the South Asian population, especially when compared to the White population. There were similar trends for OCD and ADHD within the East Asian population (Figure 2).

## DISCUSSION

This study examined whether there is a discrepancy between mental health traits and rates of mental health diagnoses for OCD, ADHD, and anxiety disorders based on ethnicity and social factors in Canada.

### Ethnic Differences in Mental Health Trait Levels

We found that East Asian and South Asian youth reported having or were reported as having significantly elevated levels of OCD traits compared to their white peers. In addition, we found that ethnic concentration was the only neighborhood-level predictor of OCD traits. This trend is similar to studies in adults that show an association between Asian Ethnic identity and several OC symptoms dimensions (Ching & Williams, 2019). Some findings from clinical samples contradict our results suggesting that OCD symptom severity does not differ based on ethnicity; however, later age of OCD symptom onset, clinical diagnosis, and treatment have been noted (J. J. Wang et al., 2020). It is possible that the increased prevalence of OCD traits within our sample reflects more developed mental illness due to a lack of diagnosis and treatment within the East and South Asian communities. These differences in findings may also be due to the nature of the sample studied. Unlike clinic studies, the present community-based sample includes East and South Asian youth with high OCD but no OCD diagnosis due to barriers to accessing care (Figure 1) (Augsberger, Yeung, Dougher, & Hahm, 2015; Islam, Khanlou, & Tamim, 2014; Yang, Rodgers, Lee, & Lê Cook, 2020).

**Figure 1:**
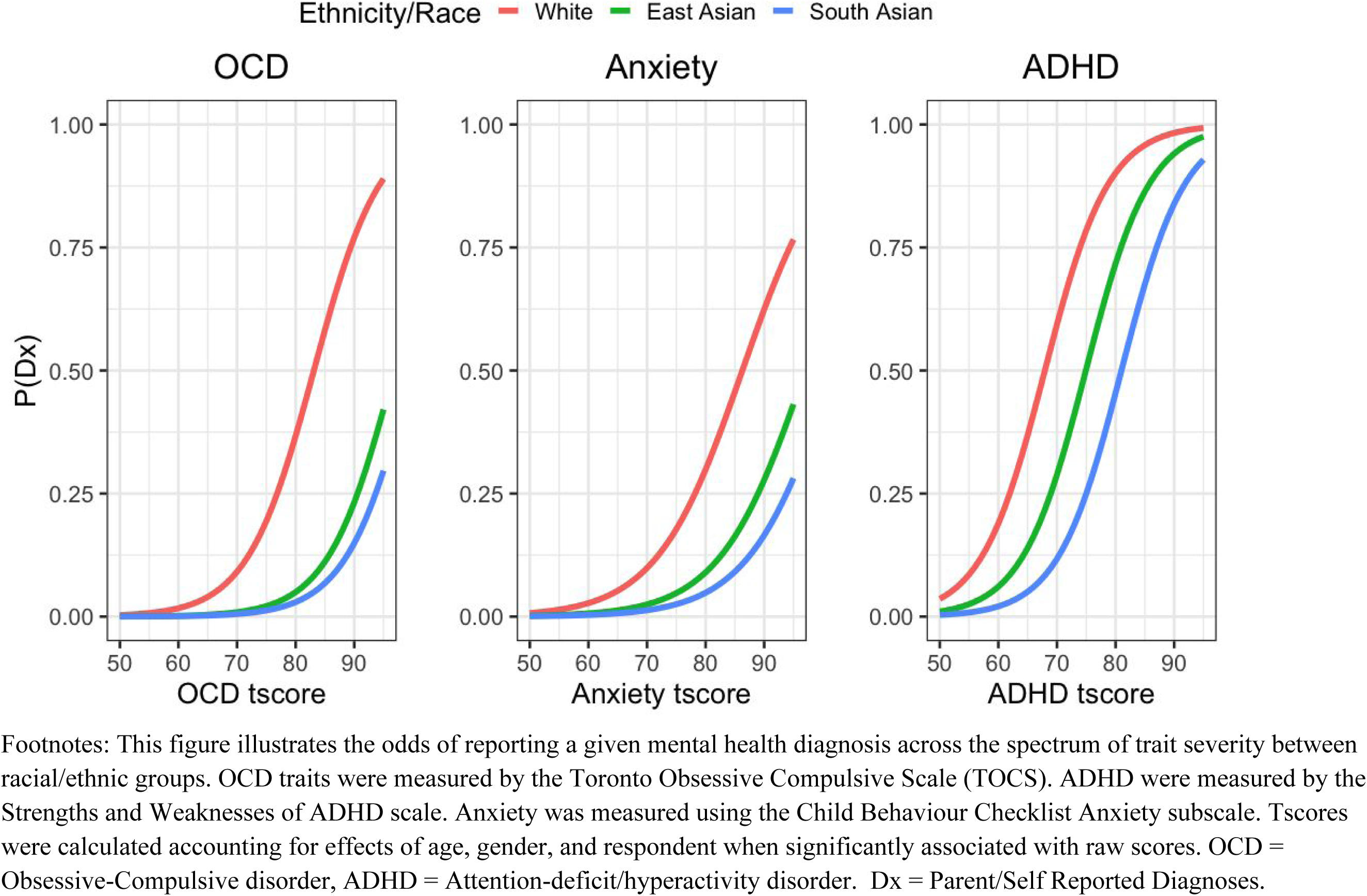
Probability of reporting OCD, ADHD, and anxiety diagnoses across the spectrum of trait severity by Racial/Ethnic Group

**Figure 2:**
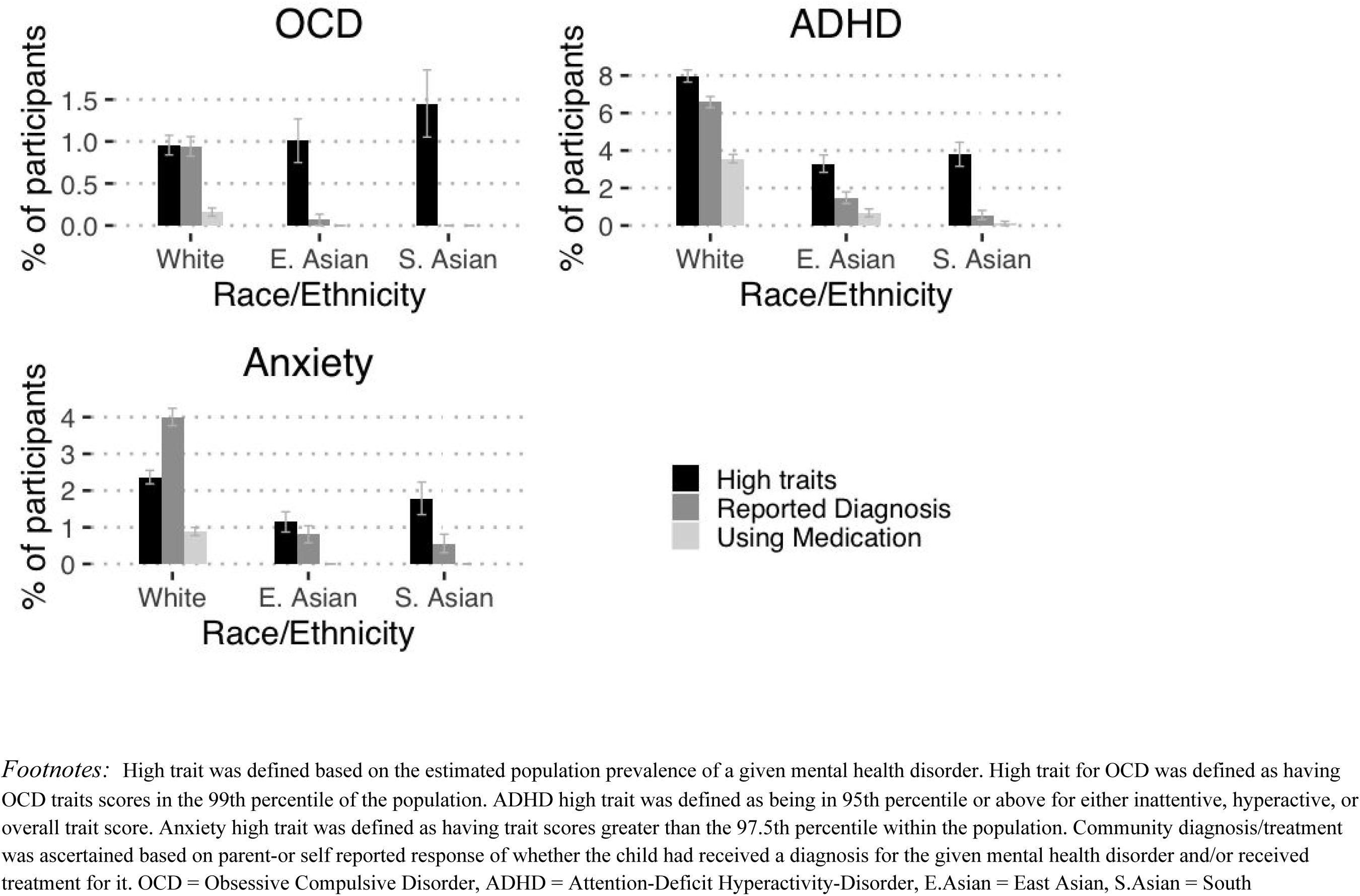
Discrepancies in Mental Health Trait levels and Rates of Diagnoses Between Racial/Ethnic Groups

We also found that East Asian and South Asian youth had significantly higher mean levels of anxiety. These results line up with studies within pediatric clinical populations within the US (Brice et al., 2015; Werner, 2020). Experiences of racism (Miller, Kim, Chen, & Alvarez, 2012) and familial/academic pressure and expectations (Saw, Berenbaum, & Okazaki, 2013) have been proposed as factors that underlie this difference. However, we also observed that increasing levels of anxiety were associated with living in a neighbourhood with increasing material deprivation. This finding suggests that socioeconomic factors may be related to risk for anxiety traits over and above the associations of ethnicity. Further work is needed to precisely elucidate the mechanisms behind these trends within Canadian youth.

We found that East Asian and South Asian youth had less reported ADHD trait levels than White youth. This result suggests that lack of ADHD trait prevalence may be one factor, in addition to potential barriers to accessing care, that may explain why numerous studies report a lower prevalence of ADHD diagnoses among Asian children (Coker et al., 2016; Getahun et al., 2013; Shi et al., 2021; Wong & Landes, 2022). Further investigation into protective factors for ADHD traits within East Asian and South Asian youth would be beneficial. We also observed that increased levels of material deprivation and decreasing socioeconomic status were associated with increased prevalence of ADHD traits. This finding was consistent with studies conducted in the UK (Russell, Ford, & Russell, 2018) suggesting that financial insecurity may be a risk factor for ADHD in youth.

### Racial/ethnic Disparities in Reporting Diagnosis and Treatment

Critically, across all mental health conditions examined in this study, we found that South Asian and East Asian youth had substantially lower odds of reporting a diagnosis and treatment than White youth at every level of trait severity. (Figure 1).The magnitude of these effects emphasizes that there are significant barriers to accessing mental health care within these racial/ethnic groups. These trends are especially concerning in the context of OCD and anxiety, where South Asians had increased trait levels relative to White youth. There are many speculative reasons behind these trends. It is well understood in adult and US populations that Asian populations have some of the lowest rates of mental health service utilization (Tiwari & Wang, 2008; Yang et al., 2020). Community, family and self-stigma may contribute to this phenomenon (Augsberger et al., 2015). One study conducted within South Asian populations residing in Ontario suggested that systemic-level, family, and community-level barriers to mental health care exist, including a lack of South Asian representation within mental health professionals, long wait times, prohibitive fees for services not covered by government healthcare coverage, and culturally insensitive treatment (Islam, Multani, Hynie, Shakya, & McKenzie, 2017). For anxiety, we also observed that having a non-English language spoken as the primary language at home was associated with decreased odds of reporting a diagnosis and/or treatment. This trend suggests that language barriers at the individual or family level may be an additional barrier to receiving a mental health diagnosis for minority ethnic children (Brisset et al., 2014). Another potential mechanism behind these trends is differences in the way that mental health traits within minority youth are interpreted by family members, teachers, guidance counselors, health care professionals and youth themselves, all of whom play key roles in the recognition of mental health disorders. Mental health care provider racial bias has also been demonstrated as a contributing factor in the misdiagnosis of ethnic and racial youth (Black Parker, McCall, Spearman-McCarthy, Rosenquist, & Cortese, 2021). Investigating the mechanisms behind these trends is complex because of the inter-relatedness of risk factors such as ethnicity and social deprivation and likely involves both structural and individual levels factors. The multifaceted nature of potential barriers to mental health diagnoses and care for East Asian and South Asian youth in Canada underscores the importance of future research to understand ethnic barriers to mental health care and their mitigation.

### Limitations

There are several limitations that should be considered within the current study. First, our main outcomes were established by self or parent-reported diagnosis as opposed to clinical diagnoses established by trained professionals. Second, given that our study utilized data from an existing genetics study we utilized participants’ maternal and paternal grandparents’ ethnicity as a proxy for participants’ race/ethnicity. That being said, we underwent multiple sensitivity analyses to ensure that the threshold for altering the definition and measurement of race/ethnicity would not alter any conclusions drawn. Future studies should aim to analyze data while asking participants for their primary self-identifying racial/ethnic group. Third, we were forced to limit our analysis to White, East Asian, and South Asian youth due to limitations in sample size. It is critical that further work is conducted with larger data sets to understand mental health disparities in different marginalized ethnic groups within Canada. Finally, we did not have any participant-level socioeconomic measures including parent immigration status and education level, all of which could have altered our results.

### Implications

Notwithstanding these limitations, there are several reasons for concern given the disparities in mental health diagnoses and care noted in this study. Most importantly, lower rates of mental health diagnoses and care may leave East Asian and South Asian youth in need of care with no access to treatment and worse eventual outcomes. Secondly, research into the treatment and etiology of mental illnesses largely focuses on cohorts of participants who have received a diagnosis. Disparities in access to diagnoses may exclude certain ethnic populations from research, further widening the research and treatment gap for historically marginalized communities.

## CONCLUSION

Within Canada, East Asian and South Asian children show significant disparities in access to mental health diagnoses and care compared to White youth, even after controlling for underlying mental health trait levels and socioeconomic factors. More work is critically needed to understand and mitigate barriers to equitable mental health care for minority ethnic youth within Canada.

**Acknowledgments:** This study was funded by the Canadian Institutes of Health Research (P.D.A.: MOP-106573; R.J.S.: MOP– 93696) and the Alberta Innovates Translational Health Chair in Child and Youth Mental Health (P.D.A.). The statistical expert on the manuscript is Dr. Annie Dupuis. The authors thank the families and participants of Spit for Science who made this study possible.

## Data Availability

The data that support the findings of this study are not openly available and are available
from the corresponding author upon reasonable request (including a study outline), subject to
review.

## SUPPORTING INFORMATION

**Table S1:** Demographics of Entire Sample

|  | Overall<br>(N=16062) |
| --- | --- |
| Residing in Canada, N (%) | 13757 (85.6%) |
| Age (years) |  |
| Mean (SD) | 11.1 (2.85) |
| Median [Min, Max] | 10.7 [6.00, 18.0] |
| Gender |  |
| female | 7935 (49.4%) |
| Ethnicity |  |
| White | 8127 (50.6%) |
| Arabic | 89 (0.6%) |
| Black | 232 (1.4%) |
| Completely Dont Know | 627 (3.9%) |
| East Asian | 1486 (9.3%) |
| Indigenous | 29 (0.2%) |
| Latin | 149 (0.9%) |
| Mixed | 2369 (14.7%) |
| Other | 230 (1.4%) |
| Pacific Islander | 7 (0.0%) |
| Partially Dont Know | 1762 (11.0%) |
| South Asian | 895 (5.6%) |
| West Asian | 60 (0.4%) |
| Respondent |  |
| self | 2900 (18.1%) |
| parent | 13162 (81.9%) |
| **Residing in Greater**<br>**Toronto Area (Urban**<br>**Subgroup** | 7435 (46.3%) |

## Description of Data Source: Spit for Science

Spit for Science was conducted in collaboration with the Ontario Science Centre which is a public space for discovery of science. Since 1969, more than 54 million people from all over Ontario, Canada, North America and world have attended. About 80% of visitors are from Ontario. About 1 million people visit each year of which about 165,000 are school children. Community access programmes assure that attendees from all walks of life are able to attend. Research Live! Is a programme that allows visitors to take part in actual scientific research and to contribute to scientific progress during their visit. Many interactive exhibits are operated by Science Centre staff; others are operated by universities and research institutes in the region. Spit for Science is an exhibit within the interactive area of the Science Centre which exhibits facts and artifacts about genes and behaviour (posters, etc). Visitors approach the exhibit to ask questions about it and are greeted by our hosts who answer their questions and invite them to participate in the study. Most participants come with a parent, but some older children come to the exhibit on their own. The Science Centre allows a 30-minute limit to participation in any project. Participation starts with a discussion of the study’s purpose followed by informed consent which has been approved by the sponsoring institution in our case The Hospital for Sick Children’s research ethics board. Research staff are trained to assess competence to consent.

Participation involves completion of rating scales to measure behavioural traits (see Methods), measurement of height and weight, completion of a brief computerized game which measures response inhibition and reaction time, and spitting into a small tube which allows us to extract DNA. Participants are rewarded with a small gift or gift certificate such as $5:00 toward movie tickets. No feedback about performance is provided and data is anonymized prior to genetic analysis. Some participants leave before finishing all aspects of the study typically because of time constraints.

